# Genetic contributions to premenstrual symptoms: revisiting the role of the *ESR1* gene

**DOI:** 10.1101/2025.01.13.25320449

**Authors:** Angelika Lahnsteiner, Esmeralda Hidalgo-Lopez, Isabel Noachtar, Tobias Hausinger, Anna Gnaiger, Katrin Griesbach, Diana Scutelnic, Angela Risch, Belinda Angela Pletzer

**Author notes:** Corresponding author: Belinda Pletzer, Department of Psychology, Centre for Cognitive Neuroscience Paris-Lodron-University Salzburg Hellbrunnerstr. 34, 5020 Salzburg, AUSTRIA.

## Abstract

Premenstrual dysphoric disorder (PMDD) and its subclinical form categorized as premenstrual syndrome (PMS), are severe mood disorders characterized by cyclical depressive symptoms, anxiety, irritability, and other functional impairments, impacting a broad range of women during the late luteal phase. The estrogen receptor 1 (*ESR1)* gene encodes the estrogen receptor alpha (ERα) which plays a critical role in mediating estrogen signaling and regulates various physiological and psychological processes. In this study, we investigated the relationship between six single nucleotide polymorphisms (SNPs) in intron 4 of the *ESR1* gene and premenstrual symptom severity, emphasizing symptom- genotype associations and SNP interactions. Results demonstrated that specific SNPs were linked to distinct symptom profiles, such as anxiety, difficulty concentrating, and sleep disturbances. Interactions between SNPs revealed both risk-enhancing and protective effects. These findings suggest that premenstrual symptoms may stem from a genotype-linked reduced sensitivity to ovarian hormones, providing a foundation for future research.

## Introduction

Premenstrual dysphoric disorder (PMDD) is a serious mood disorder characterized by the cyclical recurrence of severe depressive symptoms, anxiety, irritability, or mood lability in the final week before the onset of menses lasting up to three days after onset of menses. These symptoms are accompanied by sleep disturbances, fatigue, difficulty concentrating and a general loss of interest in everyday activities [1]. They have a significant impact on women’s day-to-day functioning and quality of life and markedly increase the risk of suicide [2]. Thus, understanding the etiology and psychobiological underpinnings of PMDD is of utmost importance.

About 3-8% of women meet the criteria for PMDD according to the fifth edition of the Diagnostic and Statistical Manual of Mental Disorders (DSM-V; [1, 3]), which requires the cyclic recurrence of at least five symptoms to be prospectively confirmed over two consecutive cycles. However, some authors argue that these criteria are overly strict since serious impairments in quality of life can already occur with fewer symptoms of significant severity. Subclinical forms of PMDD are thus often summarized as premenstrual syndrome (PMS). Given that no clear diagnostic cut-off criteria exist for PMS, prevalence rates vary across studies ranging from 9-57% [3]. Cross-cultural differences may also account for the high variability in prevalence rates [4]. For example, the highest number of PMS cases in the Global Burden of Disease Study 2019 was observed in Southeast Asia [3]. Such cultural differences may partly stem from differences in diagnostic practices. PMS/PMDD are rarely diagnosed in Western countries, and even among five Western nations, there is great variability in prescribed treatments [5]. Therefore, assessing overall symptom severity and their impact on women’s daily lives may provide a more accurate measure of the symptom burden than diagnoses, symptom counts and diagnostic cut-offs.

Accumulating evidence confirms that women with PMS or PMDD do not differ from healthy controls in their hormonal profiles throughout the menstrual cycle (reviewed in [6]). Nevertheless, hormones appear to be the trigger of psychological changes along the menstrual cycle in women with PMS/PMDD, given that the symptoms already appear during the mid-luteal cycle phase, when progesterone levels peak and estradiol levels are elevated [7]. Accordingly, it is currently hypothesized that women with PMS or PMDD exhibit increased sensitivity to hormonal fluctuations [8]. While neuroimaging studies have observed differences in brain structure, function, and neurotransmitter binding between women with PMS/PMDD and controls [9], there is currently no evidence for differences in genes related to neurotransmitter systems (reviewed in [10]).

The source of an increased reactivity to hormonal fluctuations can be suspected at the receptor level rather than the ligand level, making estrogen and progesterone receptors particularly relevant for characterizing the biological substrates of PMDD. Increased reactivity to elevated estradiol or progesterone levels may be traced back to increased sensitivity, transcriptional activity, or increased expression of these steroid hormone receptors. Given that the progesterone receptor gene is one of the transcriptional targets of estrogen receptors [11] i.e. estrogen signaling increases the expression of progesterone receptors, increased expression of estrogen receptors should result in stronger sensitivity to both estradiol and progesterone.

The estrogen receptor 1 (*ESR1*) gene is located on chromosome 6q25.1 and encodes the key transcription factor estrogen receptor alpha (ERα), which binds estrogen intracellularly and then acts in the nucleus modulating gene expression to enhance cell proliferation (reviewed in [12, 13]). Variations in *ESR1* expression have been linked to several physiological and pathological conditions, including breast cancer [14], osteoporosis [15, 16], and cardiovascular disease [17].

Single nucleotide polymorphisms (SNPs) are amongst the most common genetic variations in the human genome. Many SNPs occur in non-coding regions, where they can play crucial roles in the regulation of gene expression, particularly by influencing the binding affinity of transcription factors to their motif, or via changes in epigenetic patterns such as DNA methylation. It involves the addition of a methyl group to the cytosine base at cytosine-guanine (CpG) dinucleotides. Regulatory elements, such as gene promoters, are typically silenced when their CpG dinucleotides are highly methylated, whereas low methylation levels enable active transcription. When a SNP occurs in such a CpG dinucleotide it disrupts the CpG site leading to changes in DNA methylation patterns and therefore alterations in gene expression [18]. Another effect of SNPs is their effect on alternative splicing. Alternative splicing generates multiple transcript variants from a single gene, potentially producing proteins with different functions or regulatory properties. SNPs within intronic regions or near splice junctions can alter splicing patterns by influencing splice site selection [19]. This can result in the inclusion or exclusion of exons, e.g. leading to the production of different ERα isoforms as shown in liver [20].

Various SNPs located in intron 4 of the *ESR1* gene have been related to increased risk for breast cancer [21, 22] or increased bone mineral density [23, 24], suggesting increased estrogen signaling in minor allele carriers. So far, one study suggests that six intragenic *ESR1* SNPs increase the risk for premenstrual dysphoric disorder in a Caucasian sample [25]. These studies used PMDD diagnosis as a criterion, while focusing on symptom severity instead could relate specific symptoms to genotypes, potentially enabling the classification of PMDD subtypes based on symptom clusters.

To the best of our knowledge, previous investigations have not yet addressed whether different *ESR1* genotypes interact with each other in increasing the risk for PMDD. This question is of particular interest when addressing specific symptom clusters, as the presence or absence of minor alleles for various SNPs might either exacerbate or compensate for specific symptoms. Previous investigations have, however, reported interactions between the *ESR1* SNPs and a common SNP of the dopamine degrading enzyme Catechol-O-Methyl-Transferase (*COMT*) in modulating the risk for PMDD [25]. This interaction requires further attention given that previous evidence corroborates impairments in working memory related to premenstrual symptoms [26, 27]. Executive functions relate to fronto- striatal dopamine levels in an inverted U-shaped manner [28], and estradiol modulates central dopamine levels through multiple mechanisms (see [29] for a review). One of these mechanisms includes altered *COMT* activity with certain genotypes of the *ESR1* gene, specifically also related to one of the SNPs in the *ESR1* intron 4 [30]. Accordingly, *ESR1* genotypes also moderate the severity of cognitive decline during menopause [31]. It is thus plausible that particularly the cognitive symptoms of PMDD depend not only on estrogen sensitivity but more generally also on genes related to dopamine availability, as well as the interaction between them.

Therefore, we hypothesize that intronic *ESR1* SNPs are candidates for affecting hormone sensitivity. Here we report on the associations of six intronic SNPs of the *ESR1* gene to premenstrual symptom severity. Specifically, we explore how these SNPs interact in modulating the severity of specific symptoms. Given that recent evidence corroborates psychological symptoms as the core characteristic of PMDD, while physical symptoms did not differ significantly between women with PMS/PMDD and controls [32], we clearly distinguish between psychological and physiological symptoms in this endeavor.

## Methods

### Sample

Across four studies relating ovarian hormones to cognitive, emotional and neuroimaging parameters [33–35], 451 Caucasian female participants aged 18 to 36 years (mean age: 24.00 years, SD = 3.96) who did not use hormonal contraceptives provided both saliva samples for genetic analyses as well as information on their menstrual cycle characteristics and premenstrual symptom severity. Participants were on average highly educated with 53% having qualified for university entrance and 33% having a university degree. They were otherwise healthy with no psychiatric, endocrinological or neurological disorders. Average menstrual cycle length as calculated over at least three menstrual cycles was 29.11 (SD = 3.03). Three of these studies recruited community samples without a specific focus on severity of premenstrual symptoms to obtain a realistic estimate of the symptom distribution across the general population. In one study, 87 women were specifically recruited for suspected PMS/PMDD, though only eight of the women included in the current analysis had ever received a diagnosis of PMS/PMDD by a clinician.

### Ethics statement

All studies were approved by the University of Salzburg’s ethics committee (ethics vote numbers: GZ28/2018, GZ50/2020, GZ04/2018, GZ24/2022). All participants provided written informed consent to partake in the studies. All methods conform to the Code of Ethics of the World Medical Association (Declaration of Helsinki).

### Assessment of premenstrual symptoms

In the whole sample, premenstrual symptom severity was assessed by self-reports of participants using the premenstrual symptom screening tool (PSST). Prospective confirmation by daily ratings of severity of problems (DRSP) over two consecutive cycles was obtained from a subsample of 195 participants (mean age = 24.54 years, SD = 4.05).

#### Premenstrual symptom screening tool (PSST)

The *Premenstrual Symptom Screening Tool* (PSST, [36]) is a 20-item instrument assessing how strongly participants perceive, that symptoms like depression, anxiety, mood lability, irritability, difficulty concentrating, sleep problems, including insomnia and hypersomnia, fatigue, loss of interest in three domains, appetite changes as well as physical symptoms recur monthly in their premenstrual phase (14 items) and how strongly they perceive these symptoms impact their daily life (5 items) on a 4-point Likert scale (1 = not at all, 2 = mild, 3 = moderate, 4 = severe). The last item assesses whether participants had ever received a diagnosis of PMS/PMDD by a clinician. The PSST provides clear cut-off criteria for when a diagnosis of PMDD or PMS should be considered. A PMDD diagnosis is suspected when a total of 5 symptoms is rated as moderate to severe, including at least one of the core psychological symptoms (depression, anxiety, mood lability, irritability) rated as severe and at least one impact item rated as severe. A milder form of PMS is suspected when a total of 5 symptoms is listed as moderate to severe, including at least one of the core psychological symptoms rates as moderate to severe and at least one impact item rated as moderate to severe. The current manuscript focuses on PSST symptom strength as a continuous variable. Thus, the overall PSST score was calculated as sum over the 13 items assessing psychological symptom strength, and an Impact score was calculated as sum over the 5 items assessing the impact of PMS symptoms on daily life. Cronbach’s Alpha for the current sample was 0.92.

#### Daily rating of severity of problems (DRSP)

The *Daily Record of Severity of Problems* (DRSP, [37]) is a daily scoring sheet for patients to track common premenstrual symptoms and their impact on daily life on a 6-point Likert Scale. In its original form, the DRSP has 11 items assessing the PMS symptoms according to the DSMV. However, more detailed assessment of specific symptom clusters is possible. In the current study, we included the original 11 items as well as 5 additional items for more detailed assessment of physical symptoms (breast pain, weight gain, headaches, joint or muscle pain, nausea or abdominal pain), resulting in a total of 16 items. Thus, summary scores were computed over the 9 items assessing psychological symptoms, as well as the 5 items assessing physical symptoms. Symptom severity was assessed by calculating the percent increase from cycle days 6 to 10 (mid-follicular phase) to the 5 premenstrual days over two consecutive cycles for the overall psychological and physical symptom score respectively, as well as each individual symptom. It is recommended to consider a diagnosis of PMDD if at least 5 symptoms, one being a core psychological symptom, increase by more than 50% in the premenstrual phase. Cronbach’s Alpha for the current sample was 0.93 for psychological symptoms and 0.70 for physiological symptoms.

#### SNP genotyping using allele-specific PCRs for rs3020377 A/G, rs4680 A/G, rs3020317 C/T and rs1884051 G/A

SNP genotyping was performed with allele-specific primers using real-time PCR (Roche LightCycler) as described in our previous work [38]. In short, target regions were amplified from 10 ng genomic DNA with 1x reaction buffer, 1x EvaGreen (Jena Bioscience, Germany), 1% DMSO, 0.2 µM each of one universal and one allele-specific primer (forward and reverse primer pair) and 0.5 Units Taq Polymerase (Biozym, Austria) in a final volume of 20 µL. The reaction was pipetted two times using the allele-specific primer pair targeting each allele (e.g. A/G) in an individual reaction to compare amplification efficiency. After the first activation step at 95°C for 2 min, 95° for 15 sec, 10 sec at the annealing temperature specific for each SNP (rs3020377 = 57°C, rs1884051= 55°C, rs3020317=54°C and rs4680= 56°C), and 72°C for 5 min were repeated for 50 cycles, followed by a final amplification at 72°C for 5 min and a melt curve ranging from 65°C to 95°C with 0.5 °C/min increment. Resulting cycle threshold (Ct) values were exported to excel files. Samples were called as homozygous, if at least 5 cycles difference was obtained during the PCR. Heterozygous samples show identical Ct values, (or less than 5 cycles difference).

#### Genotyping with High Resolution Melting (HRM) Analysis for rs3020314 C/T, rs932477 A/G and rs3003917 G/A

Since we were not able to reach a sufficient separation of alleles for rs3020314, rs932477 and rs3003917 within allele-specific PCR, these three SNPs were genotyped using high resolution melting (HRM) analysis. Therefore, the target regions were amplified from 10 ng genomic DNA with 1x reaction buffer, 1x EvaGreen (Jena Bioscience, Germany), 1% DMSO, 0.2 µM each of forward and reverse primer and 0.5 Units Taq Polymerase (Biozym, Austria) in a final volume of 20 µL. The thermal protocol included an activation step at 95°C for 2 min, and 45 cycles of 95° for 15 sec, 61°C for 10 sec and 72°C for 15 sec. After a final elongation step 72°C for 5 min, a high-resolution melt curve was obtained including 95°C for 60 sec, 40°C for 60 sec, 65°C for 1 sec followed by a slow increase of temperature from 60°C to 97°C with a ramp rate of 0.07°C/sec and 15 data acquisition points per 1°C step. Data analysis was performed with the Roche Light Cycler 96 software using HRM analysis settings.

#### Genotyping with Pyrosequencing

To confirm the obtained sequencing results of HRM and allele-specific PCR, five randomly picked samples per 96 samples were genotyped using pyrosequencing. To do so, PCR products were obtained using the same PCR reaction as for HRM analysis for all seven SNPs. Pyrosequencing was performed on a PyroMark Q24 device (Qiagen,) using 20 µL PCR product and 7.5 µM sequencing primer. Detailed information can be found in the Supplement Material.

### Linkage Disequilibrium (LD) calculation

LD measures were obtained from HapMap phase 3 and visualized in UCSC genome browsers as log odds ratio (LOD) based LD map from the Central European (CEU) population [39]. Furthermore, we calculated an LD matrix of the six *ESR1* target SNPs from the 1000 Genomes project using the NCBI LDmatrix tool [40].

### Splice site prediction

To predict if the target SNPs influence alternative splicing, we used the chromosome positions and major/minor alleles for the SpliceAI-visual tool [41].

### Analysis of allele-specific gene expression influences of target SNPs and transcription factor binding

To assess the influence of genotypes on ESR1 expression, we used the expression Quantitative Trait Loci (eQTL) calculation tool within the Genotype-Tissue Expression (GTEx) Portal [42]. Binding sites for transcription factors were obtained from the University of Santa Cruz (UCSC) genome browser [43].

### Test for overlap with G-quadruplex DNA secondary structures

To determine whether the SNPs of interest are located within regions capable of forming potential DNA secondary structures, the SNP positions, along with 50 bp upstream and downstream flanking sequences, were analyzed using G4Hunter [44]. This tool predicts the likelihood of secondary structure formation based on sequence composition.

### Statistical analysis

Statistical analysis was carried out in R 4.4.1 and RStudio 2024.04.2. To establish whether subtypes of PMS/PMDD could already be identified at the symptom level, hierarchical cluster analysis was performed on PSST symptoms using the *hclust* function of the R package *cluster* [45].

To address whether *ESR1* genotypes were associated with PMS symptom strength, we performed a general linear model using the *lm* function of the *stats* package. PSST psychological symptom scores were used as dependent variable and the genotypes of the 6 SNPs of the *ESR1*, as well as their interactive effects were entered as independent variables. Genotypes were factorized with the major allele homozygous type as reference category, such that participants with one or two minor alleles were compared to the major allele homozygous type. If less than 5 cases of a certain genotype were present, groups with 1 and 2 minor alleles were merged prior to analysis. Nonsignificant interactions were backwards eliminated using the *step* function in order to allow for meaningful interpretation of main effects retained in the model.

The final model was explored for applicability to PSST diagnosis (control vs. PMS/PMDD), PSST cluster (cluster with no symptoms vs. clusters with symptoms), as well as the increase in psychological DRSP symptoms over the luteal cycle phase in the subsample with prospective symptom ratings. To relate genotypes to PSST diagnosis or cluster we performed a multiple logistic regression using the *glm* function of the *stats* package. PSST diagnosis or cluster was used as dependent variable and the genotypes associated with PSST Psychological symptoms were entered as independent variables. To relate genotypes to the DRSP symptom increase we performed linear mixed effects models using the *lme* function of the *nlme* package [46]. DRSP psychological symptoms were used as dependent variable and the genotypes associated with the PSST psychological symptoms as well as their interaction with cycle day (days to next period) were entered as independent variables.

To explore, whether the different *ESR1* genotypes were related to specific PMS symptom clusters, the final model was rerun including specific PSST and DRSP symptom scores as dependent variable. P-values were false discovery rate (FDR)-corrected for multiple comparisons using the *p.adjust* function.

To address whether PSST symptoms were related to dopamine availability, we performed a general linear model using PSST and DRSP psychological symptom scores as dependent variables and entering as independent variables the *COMT* genotypes as well as the genotypes of those *ESR1* SNPs, that have previously been related to *COMT* functioning as well as their interaction.

### Data availability

Data will be made accessible upon reasonable request.

## Results

### Diagnosis and Cluster analysis of the PSST

According to the scoring rules of the PSST, 65 women, i.e. 14% of our sample, had a suspected PMDD diagnosis, and 122, i.e. 27% of our sample, a suspected PMS diagnosis. Please note that this includes the women specifically recruited for their suspected PMS symptoms. Excluding the sample with these specific recruitment criteria 10% of women fulfilled PSST criteria for PMDD and 21% for PMS. In the subsample with prospective DRSP symptom ratings, 72% of cases with a prospective diagnosis were also detected by the PSST. However, the PSST also suspected PMS or PMDD in 40% of women, who did not fulfill the prospective DRSP diagnosis criteria.

Cluster analysis indicated three groups of women, which could be differentiated by symptom strength, but not symptom pattern. The groups roughly correspond to the 3 possible PSST diagnosis (PMDD, PMS, Control), though the overlap was not perfect. Cluster 1 included 63, i.e. 97%, of the women with a suggested PMDD diagnosis, though it also included a substantial proportion of women with subthreshold symptoms, i.e. 70 (57%) of women with a suggested PMS diagnosis. Cluster 2 included almost exclusively control women, although only 138, i.e. 52%, of women classified as controls by the PSST were included in this cluster. Cluster 3 formed an intermediate group of 51 women with a suggested PMS diagnosis (42% of PMS women) and 115 control women (44% of controls) who did not fulfill all the criteria for PMS. In summary, the cluster analysis classified a substantial proportion of PMS women as PMDD and a substantial proportion of control women as PMS. This suggests that diagnostic cut-off criteria of the PSST are set too strictly.

Across all women, the strongest symptoms were tearfulness, fatigue and physical symptoms. Only slight variations were detected across Clusters in the predominant symptom (Fig 1). Given the similarity of symptom patterns across clusters, we decided to focus subsequent analyses on symptom strength rather than diagnosis.

**Figure 1.**
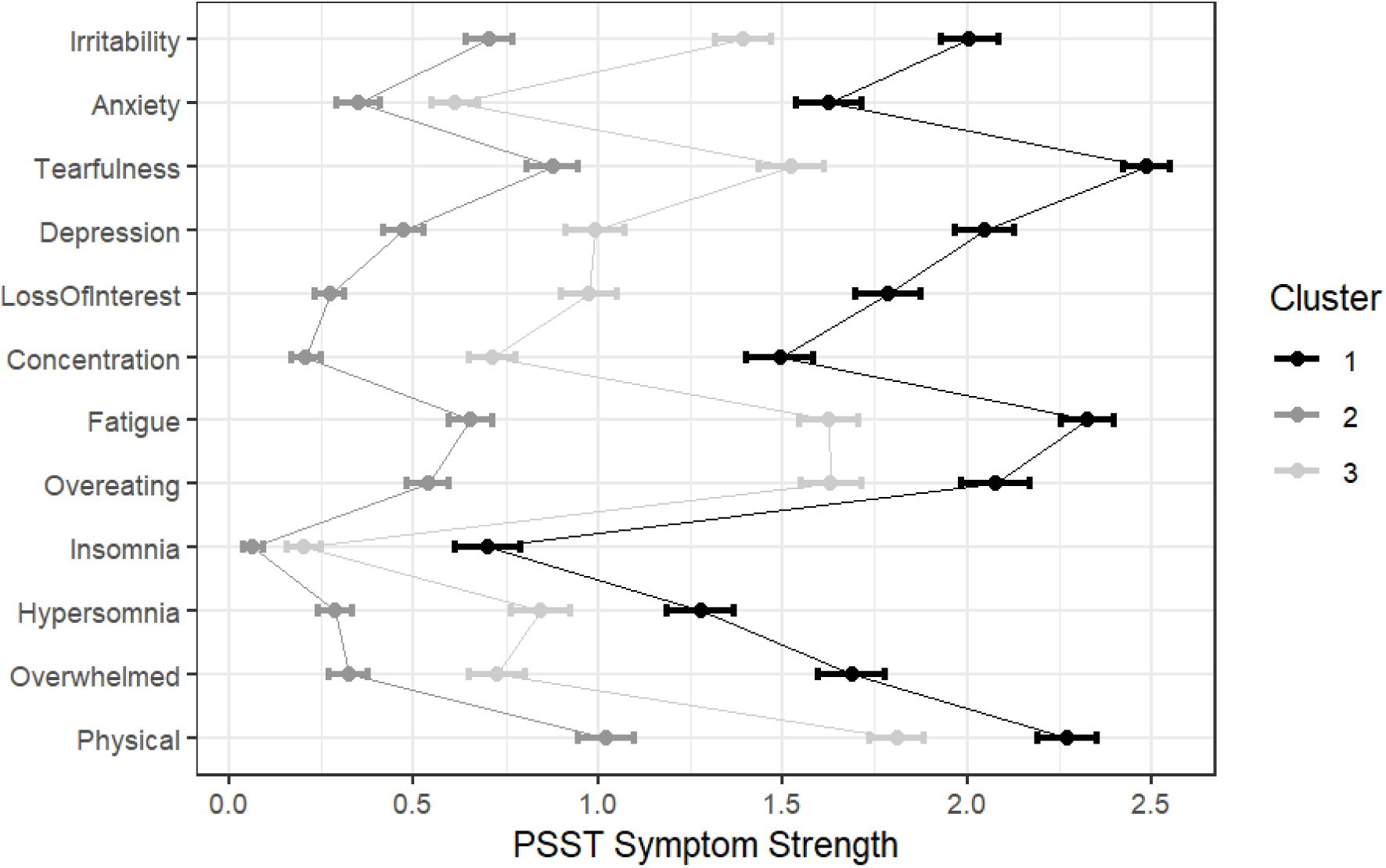
PSST symptom strength (means + 95% confidence interval) across the three clusters.

### Genotypes and association with linkage disequilibrium analysis of target SNPs

We performed genotyping of the six SNPs in *ESR1* intron 4 (rs3003917, rs3020314, rs3020377, rs3020317, rs1884051 and rs932477) and one SNP in *COMT* exon 4 (rs4680) using allele-specific PCR and high-resolution melting analysis in 451 study participants. As part of quality control, we conducted random sampling-based genotyping for all SNPs using the pyrosequencing method. All SNPs were successfully genotyped in all participants, and their genotype distributions were consistent with Hardy-Weinberg equilibrium (Table 1, chi-square test, p > 0.05).

**Table 1:**
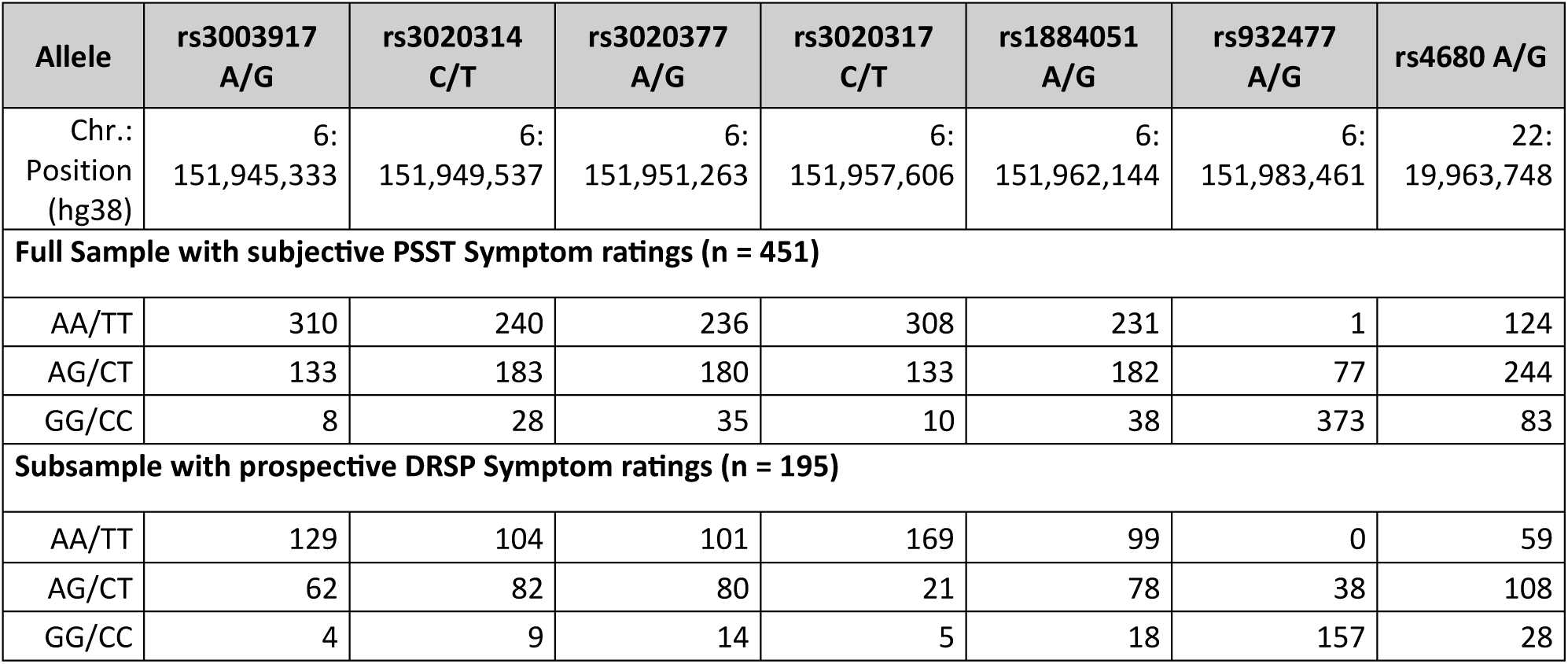
Allele frequency for the full sample and the subsample with prospective symptom ratings.

| Allele | rs3003917<br>A/G | rs3020314<br>C/T | rs3020377<br>A/G | rs3020317<br>C/T | rs1884051<br>A/G | rs932477<br>A/G | rs4680 A/G |
| --- | --- | --- | --- | --- | --- | --- | --- |
| Chr.:<br>Position<br>(hg38) | 6:<br>151,945,333 | 6:<br>151,949,537 | 6:<br>151,951,263 | 6:<br>151,957,606 | 6:<br>151,962,144 | 6:<br>151,983,461 | 22:<br>19,963,748 |
| **Full Sample with subjective PSST Symptom ratings (n = 451)** |  |  |  |  |  |  |  |
| AA/TT | 310 | 240 | 236 | 308 | 231 | 1 | 124 |
| AG/CT | 133 | 183 | 180 | 133 | 182 | 77 | 244 |
| GG/CC | 8 | 28 | 35 | 10 | 38 | 373 | 83 |
| **Subsample with prospective DRSP Symptom ratings (n = 195)** |  |  |  |  |  |  |  |
| AA/TT | 129 | 104 | 101 | 169 | 99 | 0 | 59 |
| AG/CT | 62 | 82 | 80 | 21 | 78 | 38 | 108 |
| GG/CC | 4 | 9 | 14 | 5 | 18 | 157 | 28 |

Since the six analyzed *ESR1* SNPs are located in close proximity to each other, spanning 38 kbp within intron 4, analysis of their linkage was conducted. Linkage disequilibrium (LD) describes the phenomenon where alleles at different loci on the same chromosome are inherited together more often than expected by chance due to their physical proximity. These SNPs occur in haplotype blocks which are not separated by recombination events during meiosis I. If two SNPs are in strong LD, the allele of one SNP can predict the allele of the other. Inferring LD patterns and haplotypes is crucial to identify the genetic variants that contribute to disease risk, especially for complex diseases that are influenced by multiple genetic and environmental factors [6], such as PMDD. Analysis of their linkage disequilibrium using HapMap data for the Central European (CEU) population [39] revealed that five SNPs (rs3003917, rs3020314, rs3020377, rs3020317, and rs1884051) were located within a single LD block, while one SNP, rs932477, layed outside this block (Fig. 2A). To validate these findings, we performed pairwise LD analyses using *D*^′^ and *R*^2^values derived genotyping data of the 1000 Genomes Project [40]. Stronger LD associations were indicated by higher *R*^2^ values (Fig 2B, dark red squares) and higher *D*^′^ values (Fig 2B, dark blue squares). Consistent with the Fig 2A, rs932477 showed no significant LD with any of the five SNPs in the block, as evidenced by *R*^2^ < 0.2 (Fig 2B, light red square).

**Figure 2:**
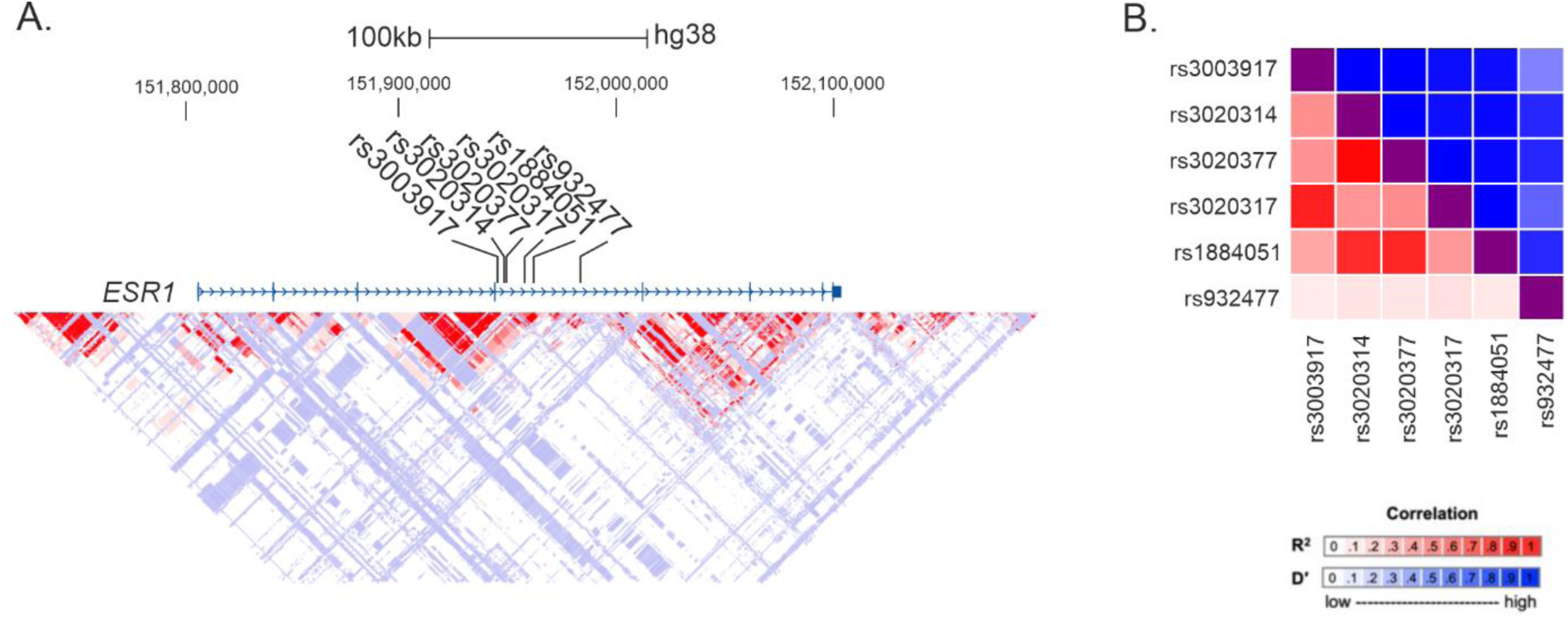
Linkage Disequilibrium (LD) Analysis. **A.** Shows an LD map inferred from the CEU population HapMap data over the whole ESR1 region. Target SNPs are indicated. Areas in dark red color indicate strong LD with high significance. Additional colors are used: White: D’<1 and no statistically significant LD (LOD < 2); light blue: D’>0.99 with low statistical significance (LOD < 2); light pink: D’<0.5 and statistical significance is high (LOD >= 2). **B.** Indicates a matrix of detailed pairwise association of target SNPs obtained from the 1000 Genomes Project.

This pattern was confirmed in our study cohort. The six SNPs within the *ESR1* gene showed strong associations with each other, i.e. participants with a minor allele on one site, were more likely to also have a minor allele at another site (all X² > 59.40, all p < 0.001). Thus, 43% of participants did not have any minor alleles. Associations were strongest for SNPs with the highest linkage equilibrium (all X² > 450), followed by SNPs with moderate linkage equilibrium (210 < X² < 310) and lowest for rs932477 (50 < X² < 160).

### Association of psychological PMS Symptoms to ESR1 SNPs

For the PSST, genotypes of all six *ESR1* SNPs were associated with premenstrual symptom severity, with two SNPs (rs1884051, rs3020317) demonstrating main effects and the remaining demonstrating two-fold interactive effects (rs3020314*rs3003917, rs3020377*rs932477). Significantly more PMS symptoms were observed in women with at least one minor G allele of rs1884051 (AG-AA: d = 0.48, t_(433)_ = 2.13, p = 0.034; GG-AA: d = 0.99, t_(433)_ = 2.83, p = 0.005; **Fig 3A**). Likewise, significantly more PMS symptoms were observed in women with 2 minor C alleles of rs3020317 (CT-TT: d = 0.03, t_(433)_ = 0.16, p = 0.872; CC-TT: d = 2.48, t_(433)_ = 2.94, p = 0.003; **Fig 3B**). However, please note that only 10 participants in the full sample were homozygous for minor C alleles of rs3020317 (compare Table 1). The main effect of rs1884051 was also confirmed when rerunning the model as multiple logistic regression using PSST diagnosis (GG-AA: d = 1.87, z = 2.32, p = 0.020) or cluster (GG-AA: d = 1.78, z = 2.27, p = 0.023) as categorical dependent variables. The rs3020317 genotype was neither associated with PSST diagnosis or cluster (all |d| < 0.69, all |z| < 1.30, all p > 0.190).

**Figure 3:**
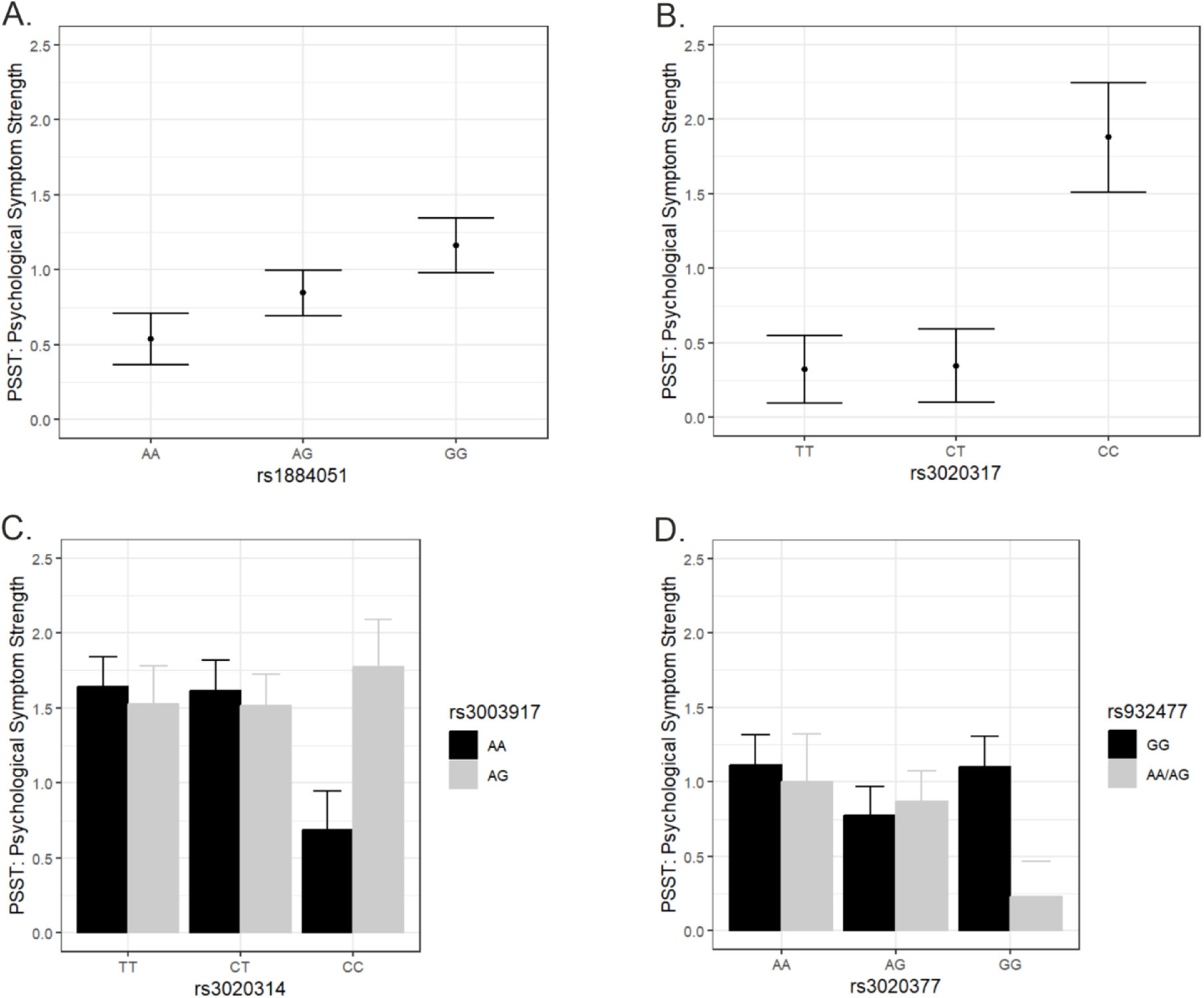
Premenstrual symptom severity across rs1884051 (A) and rs3020317 (B), as well as interactive modulation of premenstrual symptoms by rs3020314*rs3003917 (C) and rs3002377*rs932477 (D). Points and bars represent estimated marginal means adjusted for other SNPs in the model. Error bars represent standard errors. Symptom scores range from 0 (no symptoms) to 3 (severe symptoms)

Likewise, premenstrual symptom severity as assessed with the PSST was more severe in participants with minor alleles of rs3020314 and rs3020377. However, these effects were moderated by the genotypes of rs3003917 and rs932477 respectively. Severe psychological symptoms in participants with minor alleles of rs3020314 were only observed if they also had at least one minor allele for rs3003917 (CC-TT*AG-AA: d = 1.92, t_(433)_ = 3.10, p = 0.002; CC-TT*GG-AA: d = 2.42, t_(433)_ = 2.01, p = 0.045; **Fig 3C**).

In contrast, severe psychological symptoms in participants with minor alleles of rs3020377 were restricted to participants with the common genotype GG for rs932477 (GG-AA*AA/AG-GG: d = -1.22, t_(433)_ = -2.23, p = 0.026; **Fig 3D**). The interaction between rs3020314 and rs3020377 was also confirmed when rerunning the model as multiple logistic regression using PSST diagnosis (CC-TT*GG-AA: d = 4.08, z = 2.62, p = 0.009) or cluster (CC-TT*GG-AA: d = 3.90, z = 2.38, p = 0.017) as categorical dependent variables. The interaction between rs3003917 and rs932477 was only by trend associated with PSST diagnosis (GG-AA*AA/AG-GG: d = -2.11, z = -1.71, p = 0.087), but was also significantly associated with cluster (GG-AA*AA/AG-GG: d = -3.27, z = 2.58, p = 0.010).

In summary, PSST ratings suggest that patients are at risk for developing more severe premenstrual symptoms if they have (i) at least one minor allele of rs1884051, (ii) two minor alleles of rs3020317, (iii) minor alleles of both rs3020314 and rs3020917, (iv) two minor alleles of rs3020377, but no minor allele of rs932477.

In the subsample with prospective symptom ratings, we explored the relationship of genotypes to the development of absolute DRSP scores over the luteal cycle phase using a linear mixed effects model. Results revealed a significantly stronger increase in psychological symptoms over cycle days in women with one minor allele of rs1884051 (cycle_day*AG-AA: d = 0.10, t_(6013)_ = 2.69, p = 0.007) or rs3020317 (cycle_day*CT-TT: d = 0.13, t_(6013)_ = 2.35, p = 0.018). Furthermore, the model confirmed a stronger increase in psychological symptoms over cycle days in women with minor alleles of both rs3020314 and rs3020917 (cycle_day*CC-TT*AG/GG-AA: d = 0.45, t_(6013)_ = 2.80, p = 0.005) or two minor allels of rs3020377, but no minor allele of rs932477 (cycle_day*GG-AA*AA/AG-GG: d = -0.45, t_(6013)_ = -3.98, p < 0.001).

### Symptom clusters related to specific genotypes

For the PSST, differences between ESR1 genotypes were most pronounced for anxiety, and also extended to loss of interest, concentration and sleeping problems. Interestingly, the different SNPs related to distinct symptom patterns. While minor alleles of rs1884051 were associated with psychological symptoms, like anxiety, rs3020317 was associated with cognitive symptoms, like trouble concentrating and insomnia (compare **Table 2**). Symptoms related to rs1884051, i.e. anxiety, were also related to the interactive effects of rs3020377 and rs932477. Symptoms related to rs3020317, i.e. troubles concentrating, were also related to the interactive effects of rs3020314 and rs303197.

**Table 2:** Standardized effect sizes for associations between *ESR1* genotypes and specific premenstrual symptoms. Standard errors are listed in brackets. *p_FDR_ < 0.05, **p_FDR_ < 0.01, ***p_FDR_ < 0.001

|  | rs1884051[GG-AA] | rs3020317[CC-TT] | rs3020314[CC-TT]*<br>rs3003917[AG-AA] | rs3020377[GG-AA]*<br>rs932477[AG-AA] |
| --- | --- | --- | --- | --- |
| Irritability | 0.63 (0.36) | 0.55 (0.41) | 1.07 (0.63) | -0.79 (0.56) |
| **Anxiety** | **1.12 (0.34)\*\*** | 1.39 (0.83) | 1.40 (0.61) | **-2.79 (0.54)\*\*\*** |
| Tearfulness | 0.58 (0.36) | 0.45 (0.86) | 0.86 (0.63) | -0.80 (0.56) |
| Depressivity | 0.53 (0.36) | 1.77 (0.86) | 1.23 (0.63) | -1.21 (0.56) |
| **Loss of interest** | 0.82 (0.36) | **2.19 (0.85)\*** | **1.91 (0.63)\*** | -0.22 (0.56) |
| **Concentration** | 0.21 (0.35) | **3.11 (0.84)\*\*\*** | **1.70 (0.62)\*** | -0.79 (0.55) |
| Fatigue | 0.32 (0.36) | 1.98 (0.86) | 0.76 (0.63) | -1.12 (0.56) |
| Overeating | 0.71 (0.36) | 0.48 (0.86) | 0.70 (0.63) | -0.55 (0.56) |
| **Insomnia** | 0.41 (0.36) | **2.55 (0.85)\*\*** | 0.99 (0.63) | -0.30 (0.55) |
| **Hypersomnia** | 0.85 (0.35) | **1.16 (0.85)\*** | **1.51 (0.53)\*** | -0.21 (0.55) |
| Overwhelmed | 0.62 (0.36) | 0.98 (0.86) | 0.56 (0.64) | -0.86 (0.56) |

### Associations of *ESR1* genotypes to physical symptoms

In the PSST, physical symptoms were addressed by one item, which was not related to any of the ESR1 SNPs (all |d| < 0.98, all |t| < 1.57, all p > 0.11). The DRSP assessed breast tenderness, weight gain, headaches, nausea and joint pain. A significant association of *ESR1* genotypes was identified for headaches, but none of the other physical symptoms (all |d| < 0.90, all |t| < 1.95, all p > 0.05). Interestingly for headaches, there were main effects for both rs3020377 (AG-AA: d = 1.54, t_(175)_ = 3.23, p_FDR_ = 0.005; GG-AA: d = 0.42, t_(175)_ = 0.68, p = 0.500; **Fig 4A**) and rs3020314 (CT-TT: d = -1.89, t_(175)_ = -4.25, p_FDR_ < 0.001; CC-TT: d = -0.57, t_(175)_ = -0.95, p = 0.340; **Fig 4B**) that were not modulated by other genotypes.

**Figure 4:**
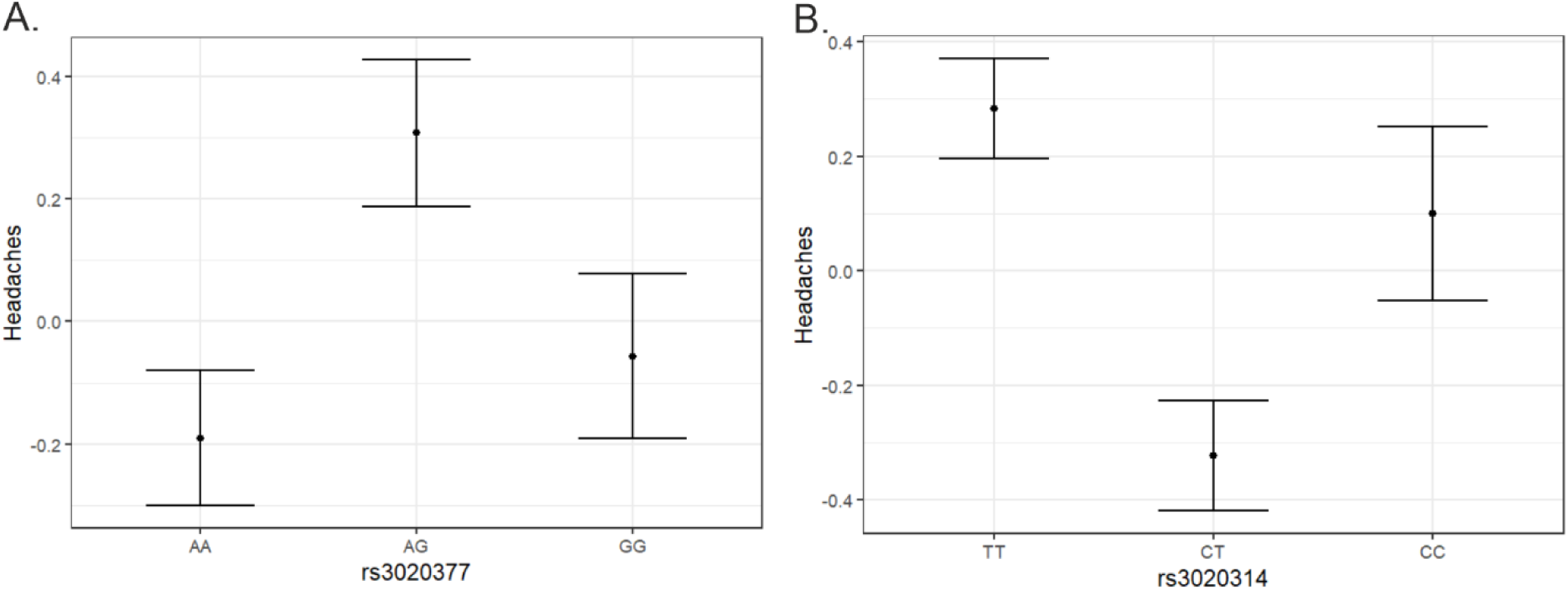
Premenstrual increase in headache severity across rs3020377 and rs3020314. Points represent estimated marginal means adjusted for other SNPs in the model. Error bars represent standard errors.

### Interactive modulation of PMS symptoms by *ESR1* and *COMT*

For the PSST, there was a significant interaction between the genotype of rs3020377 and the *COMT* rs4680 genotype (GG-AA*GG-AA: d = -1.08, t = -1.96, p = 0.05; **Fig 5A**). This interaction was even more pronounced for the DRSP (cycle_day*AG-AA*GG-AA: d = -0.60, t_(6044)_ = -5.26, p < 0.001; GG-AA*GG-AA: d = -0.60, t_(6044)_ = -4.33, p < 0.001). For participants homozygous for major alleles of rs3020377, who are presumed to have low estrogen signaling, there was a u-shaped relationship to the *COMT* genotype, such that the lowest psychological PMS Symptoms were observed in subjects heterozygous for rs4680, i.e. with moderate *COMT* activity. For participants with one minor allele of rs3020377, who are presumed to have moderate estrogen signaling, this relationship was reversed, with lowest severity of psychological PMS symptoms for participants with low *COMT* activity.

**Figure 5:**
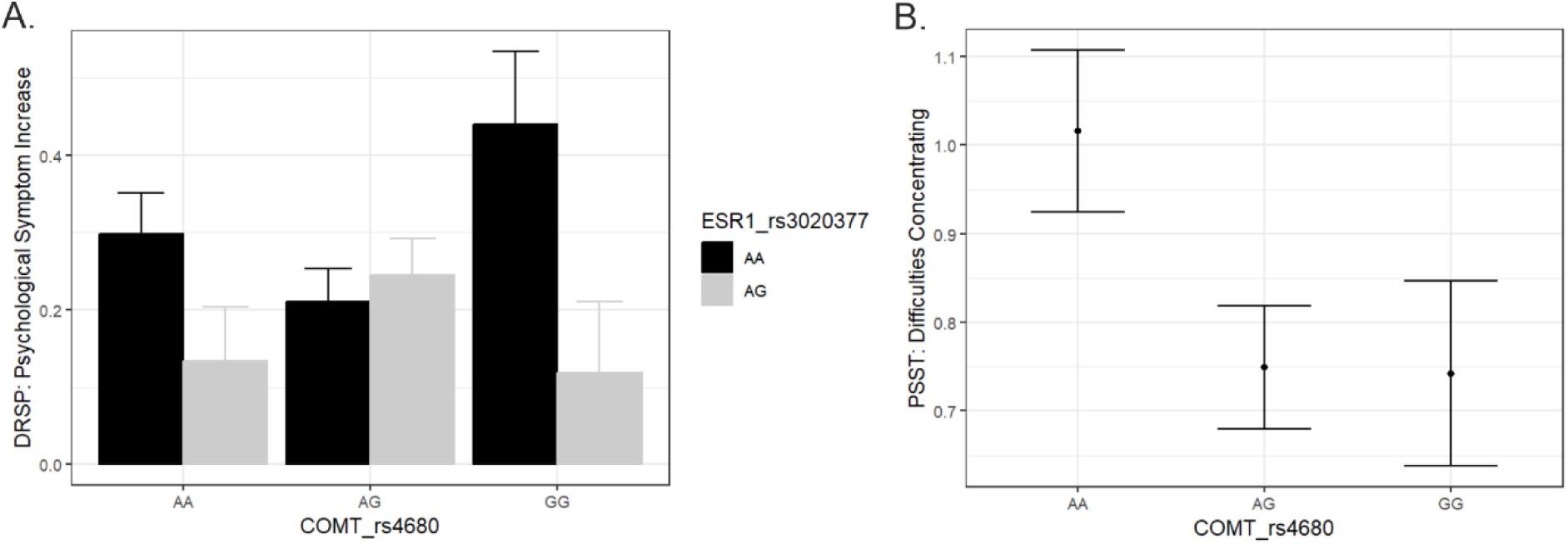
Association of COMT-Genotype to PMS Symptoms. (A) interactive modulation of psychological symptoms by ESR1 SNP rs3020377 and COMT SNP rs4680. (B) Association of COMT Genotype to Difficulties concentrating.

Exploring whether this association was driven by difficulties concentrating, the interaction was not confirmed, but a main effect of *COMT* emerged (AG-AA: d = -0.30, t_(446)_ = -2.89, p = 0.006; GG-AA: d = - 0.31, t_(446)_ = -2.16, p = 0.031; **Fig 5B**) with lowest difficulties concentrating for subjects heterozygous for rs4680, i.e. with moderate COMT activity. This effect was, however, not confirmed for the DRSP (AG- AA: d = -0.11, t_(180)_ = -0.65, p = 0.51; GG-AA: d = -0.10, t_(180)_ = -0.40, p = 0.690). We also investigated a potential link to menstrual cycle length. While *ESR1* genotypes did not show any association (data not shown here), the *COMT* genotype was significantly associated with cycle duration, such that a higher number of G-Alleles was related to longer cycles (GG – AA: d = 0.45, t(442) = 3.20, p = 0.002; Supplement Fig. 1). Menstrual cycle irregularity was not related to *ESR1* or *COMT* genotypes.

### Target SNP alleles are associated with differential expression in different tissues

Introns represent the non-coding portions of genes. Although historically regarded as “junk DNA,” it is now well established that these regions harbor important regulatory elements that control gene expression [47]. SNPs have been shown to influence gene expression through various mechanisms: (i) by altering splice sites, thereby generating new transcript variants, and (ii) by modifying transcription factor (TF) binding motifs, which can affect the binding affinity of TFs. To investigate these potential mechanisms, we analyzed splice sites using an alternative splice site prediction tool [41]. This analysis did not reveal any impact of the target SNPs in *ESR1* on alternative splicing. In contrast, analysis of TF binding motifs inferred from the University of Santa Cruz (UCSC) genome browser [43] overlapping with or adjacent to the SNPs demonstrated that all SNPs either overlapped or were in close proximity (within 10 bp) of binding sites (Supplement Fig. 2). SNPs can alter the sequence of a transcription factor binding motif either directly or indirectly over longer distances if the SNP is located in a region capable of forming a DNA secondary structure, such as a G-quadruplex (G4s) [48]. G4s play multiple critical roles in the genome and are known as transcription factor binding hubs [49]. SNPs that disrupt the motif necessary for G4 folding can also alter transcription patterns [50]. The effects of these changes can extend over wider ranges of approximately 20–60 bp, depending on the length of the motif forming the secondary structure and the SNP’s position within it [50]. Nevertheless, none of the investigated SNPs directly overlapped with a G4-forming motif, as assessed using the G4Hunter tool (data not shown here) [44]. This suggests that these SNPs are unlikely to impact DNA secondary structure formation.

To assess whether *ESR1* gene expression varies across tissues based on genotype, publicly available expression and genotype data from the Genotype-Tissue Expression (GTEx) Portal were analyzed [42]. Notably, while many SNPs and tissues did not show any significant association, rs3020317 and rs932477 were associated with significant changes in *ESR1* transcription in specific tissues: rs932477 in the aorta and ovary, and both rs3020317 and rs932477 in the hypothalamus (Supplementary Table 1 and Supplement Fig 3). The minor allele A of rs932477 was associated with higher *ESR1* expression in these tissues, whereas the minor allele C of rs3020317 was associated with lower expression compared to heterozygous (CT) samples. It is noteworthy that the overall expression of *ESR1* was very low in brain tissue and arteries, and thus the presence of the minor allele resulted in only a slight increase in *ESR1* expression (Supplement Fig 4).

## Discussion

The current study addressed the question, whether six SNPs located in intron 4 of the *ESR1* gene were related to premenstrual symptoms severity. Specifically, we asked, which symptoms were associated with which genotypes and whether interactions among SNPs modulated premenstrual symptoms. Replicating the study of Huo et al. [25] we found that all six SNPs related to premenstrual symptoms. However, our study improves upon this by leveraging a larger sample size and focusing on premenstrual symptom severity rather than diagnosis. This allowed us to: (i) provide first evidence for potential interactions between SNPs in regulating premenstrual symptoms and (ii) link genotypes from different haplotype blocks to distinct symptom patterns. In addition, two of the *ESR1* SNPs were also associated with headaches, while concentration issues were linked to *COMT* genotypes.

Regarding the associations between *ESR1* SNPs and premenstrual symptom strength, we observed stronger premenstrual symptoms with minor alleles of rs1884051 and rs3020317, as well as interactive effects between rs3020314 and rs3003917 on the one hand, and rs3020377 and rs932477 on the other hand. It is important to note that no interactions emerged between SNPs with strong linkage disequilibrium (LD). Rather, multiple regression analysis revealed which of the SNPs with the strongest LD showed the strongest association with premenstrual symptoms. The interactive effects demonstrate two different possible scenarios. While minor alleles of rs3020314 and rs3003917 appear to reinforce each other’s effects on premenstrual symptom strength, minor alleles of rs3020377 and rs932477 appear to counteract each other’s effects on premenstrual symptom strength. These observations suggest that the role of *ESR1* SNPs is more complex than previously assumed. While some SNPs may reduce each other’s contribution to the risk of developing PMDD, others may in combination accumulate the risk. A potential protective or counterbalancing effect may offer an evolutionary explanation for the LD patterns observed among SNPs in the current study. For example, in the case of rs3020317, the most severe PMDD symptoms were observed when no minor allele of rs3003917 was present. However, due to the strong LD between these two SNPs, only two participants in the sample fulfilled this criterion, such that the interactive effect between these two SNPs did not reach statistical significance. An interesting avenue for future even larger-scale studies would be to explore whether participants with minor alleles of only one SNP from the same haplotype block exhibit the most severe premenstrual symptoms. This is of particular importance, as prolonged lifetime exposure to estrogen caused by younger age of the first period is associated with an increased risk of developing breast cancer [51] and expression of *ESR1* may be at least partly influenced by the same SNPs which are associated with PMS symptoms.

Regarding symptom specificity, it is an interesting observation, that the strongest associations were observed for anxiety, despite irritability and mood swings being the core symptoms of PMS [32] (compare also Figure 1). In fact, associations between minor alleles and anxiety were twice as high as those between minor alleles and other psychological symptoms. It has previously been demonstrated that the anxiogenic effects of estrogens are mediated via ERα, while the anxiolytic effects of estrogens are mediated via ERβ (reviewed in [52]). Thus, while the postulated hormone sensitivity among women with PMDD is in fact reflected in the SNPs investigated in the current study, they only represent one part of the picture, and further evaluations are necessary to elucidate the genetic underpinnings for other psychological symptoms like irritability, depression, or mood swings. Given that even among the six SNPs investigated, some showed stronger associations with anxiety, while others exhibited stronger associations with cognitive and motivational symptoms like loss of interest and trouble concentrating, it is possible that other loci within the *ESR1* gene of the *ESR2* gene contribute to additional symptoms.

Importantly, the observation that SNPs from different haplotype blocks relate to different symptom clusters highlights the heterogeneity of the syndrome, which extends beyond what is identifiable via cluster analysis of questionnaire items. Gaining a deeper understanding of genotypes underlying specific symptoms could facilitate future development of targeted treatments for various subtypes of PMDD.

In line with the diagnostic validity of the PMDD diagnosis [32], the modulation of premenstrual symptoms by *ESR1* appears to be restricted to certain psychological symptoms, while physical symptoms were not related to *ESR1* genotypes. An exception to this general pattern are headaches, which were also related to two *ESR1* SNPs located in intron 4 in the current investigation. However, they appear to follow a different pattern of associations with u-shaped and inverted u-shaped rather than linearly increasing relationships with higher number of minor alleles. It is particularly striking that two SNPs in strong linkage equilibrium exhibit opposite association patterns with headaches. While headaches increase with one minor allele of rs3020377, they decrease with one minor allele of rs3020314. This supports the idea that the linkage of minor alleles may in some cases serve a protective effect. Two linked biological mechanisms may be candidates for a common modulation of psychological symptoms and headaches by *ESR1* genotypes. First, alterations in the serotonergic system have been linked to mental health symptoms like depression and anxiety [53] and pain perception, specifically headaches and migraines [54]. Second, vascular health, more specifically vasodilation and vasoconstriction are linked to stress and anxiety on the one hand [55], but headaches and migraines on the other hand [56]. Interestingly, serotonin can have vasoconstrictive or vasorelaxant effects depending on the predominant receptor type in certain brain areas [57]. Estradiol is generally considered a vasorelaxant hormone. Specifically, the *ESR1* SNP rs3020377 was previously related to endothelium-independent vasodilation [58].However, recent evidence suggests that, like serotonin, estradiol may also exert vasoconstrictive effects under certain conditions [59]. Thus, altered estrogen signaling due to different *ESR1* genotypes may affect psychological symptoms and headaches via interactions with the serotonergic system or vascular processes. In this context it is of particular interest that the *ESR1* SNPs investigated in the current study appear to affect *ESR1* expression, particularly in blood vessels like the aorta (compare Supplementary material).

Evaluation of the target SNPs revealed significant associations between rs3020317 and rs932477 and *ESR1* gene expression in specific tissues. Specifically, rs932477 was linked to increased *ESR1* expression in the aorta and ovary, while both rs3020317 and rs932477 influenced expression levels in the hypothalamus. The minor allele A of rs932477 was consistently associated with elevated ESR1 expression, whereas the minor allele C of rs3020317 correlated with reduced expression relative to heterozygous (CT) samples. Given that minor alleles of rs3020317 were associated with increased premenstrual symptoms, but minor alleles of rs932477 were associated with reduced premenstrual symptoms, this raises the question whether the cause of premenstrual symptoms indeed lies in increased sensitivity to ovarian hormones. The pattern of associations between these SNPs and premenstrual symptoms on the one hand and gene expression on the other hand, suggests that instead of an increased hormone sensitivity, a reduced hormone sensitivity might be associated with premenstrual symptoms. It has repeatedly been demonstrated that the healthy female brain adapts – primarily via connectivity changes – to ovarian hormone fluctuations along the menstrual cycle [60–62]. It has also been demonstrated that in the case of cognitive tasks, these adaptations occur in the absence of cognitive changes and thus serve to uphold the status quo in changing hormonal milieus [35, 60]. It is possible that the same applies to emotion regulation and mental health. If estrogen receptor expression is lower due to certain genotypes, it is possible that the brain does not respond as strongly to ovarian hormones, limiting the connectivity adaptations, that serve to balance the effects of ovarian hormone fluctuations across multiple systems.

Additional analyses revealed that *ESR1* SNPs not only influence each other’s effects on premenstrual symptoms but also modulate their association with the *COMT* rs4680 polymorphism. This polymorphism results in an amino-acid change (Val158Met) directly affecting the enzymatic activity of COMT. Thus, the non-linear interaction between *ESR1* SNP rs3020377 and *COMT* SNP rs4680 is likely attributable to the estrogenic modulation of dopamine signaling [29] and may explain why previous investigations failed to demonstrate a direct association between premenstrual symptoms and polymorphisms in the *COMT* gene or dopamine receptor genes [25, 63]. These findings suggest that dopaminergic contributions to premenstrual symptom strength are moderated by estrogen sensitivity. However, this does not apply for concentration difficulties, for which a main effect of the COMT Val158Met polymorphism was observed. However, women with two A alleles — Met carriers with lower *COMT* activity and thus higher central dopamine levels — reported the highest concentrating difficulties. It is possible that women with an overall high level of executive functioning experience a premenstrual impairment in concentration due to psychological symptoms as a stronger limitation than women with generally lower executive control. Contrary to this assumption, previous studies have demonstrated a working memory impairment in women with PMDD [26, 27], although it is currently unclear whether the impairment represents a trait or is restricted to the premenstrual phase.

Finally, we explored associations of the investigated SNPs to parameters beyond premenstrual symptom severity and observed that the *COMT* rs4680 polymorphism was also related to menstrual cycle duration. Specifically, G alleles, which are associated with higher enzyme activity and thus lower dopamine levels related to longer menstrual cycles. It is possible that this association reflects an indirect effect of dopamine on GnRH neurons via the inhibition of prolactin release [64].

In summary, we found SNPs in intron 4 of the *ESR1* gene associated with premenstrual symptom severity, particularly anxiety symptoms, difficulty concentrating and sleeping problems. Importantly, different SNPs related to different problems and SNPs interacted with each other in their association to symptom severity. Comparing the associations between the studied SNPs and premenstrual symptoms to the associations between the studied SNPs and *ESR1* expression, opened an interesting new avenue for interpreting premenstrual symptoms. Future studies may want to consider the possibility of an insensitivity to ovarian hormones as a potential mechanism behind the development of premenstrual symptoms, as well as associations between premenstrual symptoms and breast cancer risk.

## Acknowledgments

We thank Julia Kinzelman, Alina Panzik, Laura Smida, Livia Rauter, Magdalena Siebers, Patricia Gruschwitz, Patricia Werlein and Rebecca Wagenbrenner for their help with data acquisition and sample preparation and all participants for their time and willingness to contribute to this study.

