## Supplement_File for "Genetic contributions to premenstrual symptoms: revisiting the role of the *ESR1* gene"

### 1. Supplement Material

#### 1.1. Allele-specific primers

| SNP | Position (hg38) | Primer-ID | Sequence 5´-3´ | Amplicon length [bp] |
|---|---|---|---|---|
| rs4680 A/G | chr22:19963748 | F-4680 | GGGCCTACTGTGGCTACTC | 146 |
| | | R-4 680A | CATGCACACCTTGTCCTT*T*A*T | |
| | | R-4 680G | CATGCACACCTTGTCCTT*T*A*C | |
| rs3020377 A/G | chr6:151951263 | F-3020377A | CAATGTCGTTTTCCTACAGA*A*G*A | 156 |
| | | F-3020377G/2 | CAATGTCGTTTTCCTACAGA*T*G*G | |
| | | R-3020377 | GGCCAATGGGTCATATCTTTC | |
| rs3020317 C/T | chr6:151957606 | F-3020317C | GCCAGTTTTCGGAGTTAA*A*A *C | 234 |
| | | F-3020317T | GCCAGTTTTCGGAGTTAA*A*A *T | |
| | | R-3020317 | GTTATGCCAAGGCTTTATCTTG | |
| rs1884051 G/A | chr6:151962144 | F-1884051A | AGGATCAAGAGCTTCTG*T*C *A | 159 |
| | | F-1884051G | AGGATCAAGAGCTTCTG*T*C *G | |
| | | R-1884051 | TCCTAAAGGGCCTCAGGATAAG | |

Green = allele-specific base
Red = mismatched base to increase specificity. For more details see Heissl et al. 2017 of the reference list in the main text.
* = Phosphorothioate bond (PTO)

#### 1.2. High resolution melting (HRM) analysis primers

| SNP | Position (hg38) | Primer-ID | Sequence 5´-3´ | Amplicon length [bp] |
|---|---|---|---|---|
| rs3003917 G/A | chr6:151945333 | F-3003917/HRM | ATCCAACCTGGTGTATGCTTAG | 110 |
| | | R-3003917/HRM | ACTTCAGCCACTGTGTCTTTC | |
| rs3020314 C/T | chr6:151949537 | F-3020314/HRM | CATCCTGGAGAGATGACAGAAG | 101 |
| | | R-3020314/HRM | CGATTCCAGCCCAATCCTAAG | |
| rs932477 A/G | chr6:151983461 | F-932477/HRM | AGCTCAAGTAGCTTGGTGTC | 105 |
| | | R-932477/HRM | TAAGCTTCAGGAAGCCACAG | |

##### 1.3. Pyrosequencing primers

| SNP | Position (hg38) | Primer-ID | Sequence 5´-3´ | Amplicon length |
|---|---|---|---|---|
| rs3020377 A/G (*ESR1*) | chr6:151951263 | F-3020377 | TTCATCCTGGTATGCTGTGTGAG | 84 |
| | | R-3020377-bio | GTTCGAATGAAACCGAAACAACT | |
| | | S-3020377 | TGTCGTTTTCCTACAGAT | |
| rs3020314 C/T (*ESR1*) | chr6:151949537 | F-3020314PS | TTGGCAAGGTAATGAAAGATAACA | 111 |
| | | R-3020314PS-bio | AGTGGGTGCAAAATCTAAATGG | |
| | | S-3020314 | CTGGAGAGATGACAGAAG | |
| rs3020317 C/T (*ESR1*) | chr6:151957606 | F-3020317PS | ATGCCTTTCTCTTCAAACAATCTT | 165 |
| | | R-3020317PS-bio | ACTTAAATGTAGCCCCAAATCACT | |
| | | S-3020317 | CCAGTTTTCGGAGTTAAT | |
| rs1884051 G/A (*ESR1*) | chr6:151962144 | F-1884051PS | TCCACTAGCTTGCTGCTTGTCT | 82 |
| | | R-1884051PS-bio | ACTCCCTTGAAGCAGCAGAATG | |
| | | S-1884051 | ATCAAGAGCTTCTGCC | |
| rs3003917 G/A (*ESR1*) | chr6:151945333 | F-3003917PS-bio | TTATCCAACCTGGTGTATGCTTAG | 111 |
| | | R-3003917PS | CTTCAGCCACTGTGTCTTTCTAAA | |
| | | S-3003917 | GCCTTTCTCTTTCTCCTA | |
| rs932477 A/G (*ESR1*) | chr6:151983461 | F-932477PS-bio | GGCCCATTTTTCAGAAAGGA | 50 |
| | | R-932477PS | TTGGCACACCCAGGATCAC | |
| | | S-932477 | GCACACCCAGGATCAC | |
| rs4680 A/G (*COMT*) | chr22:19963748 | F-4680-bio | CCAGCGGATGGTGGATTTC | 84 |
| | | R-4680 | ACAGCCGGCCCTTTTTCC | |
| | | S-4680 | GCACACCTTGTCCTTC | |

#### 2. Supplementary Figures

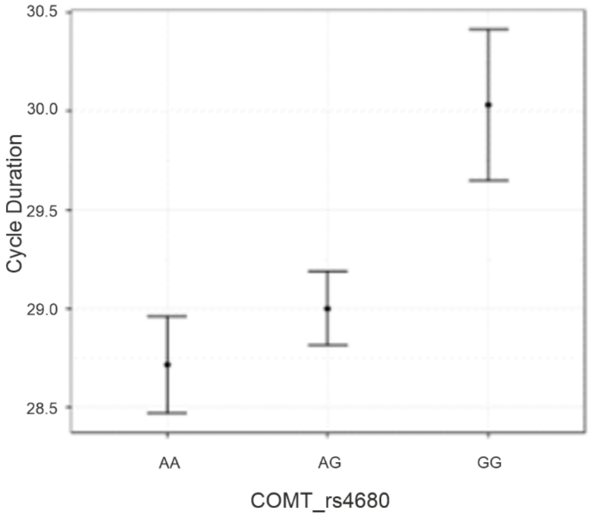

**Supplement Figure 1. Menstrual cycle characteristics.** Association of *COMT* Genotype to menstrual cycle duration. Error bars represent standard errors.

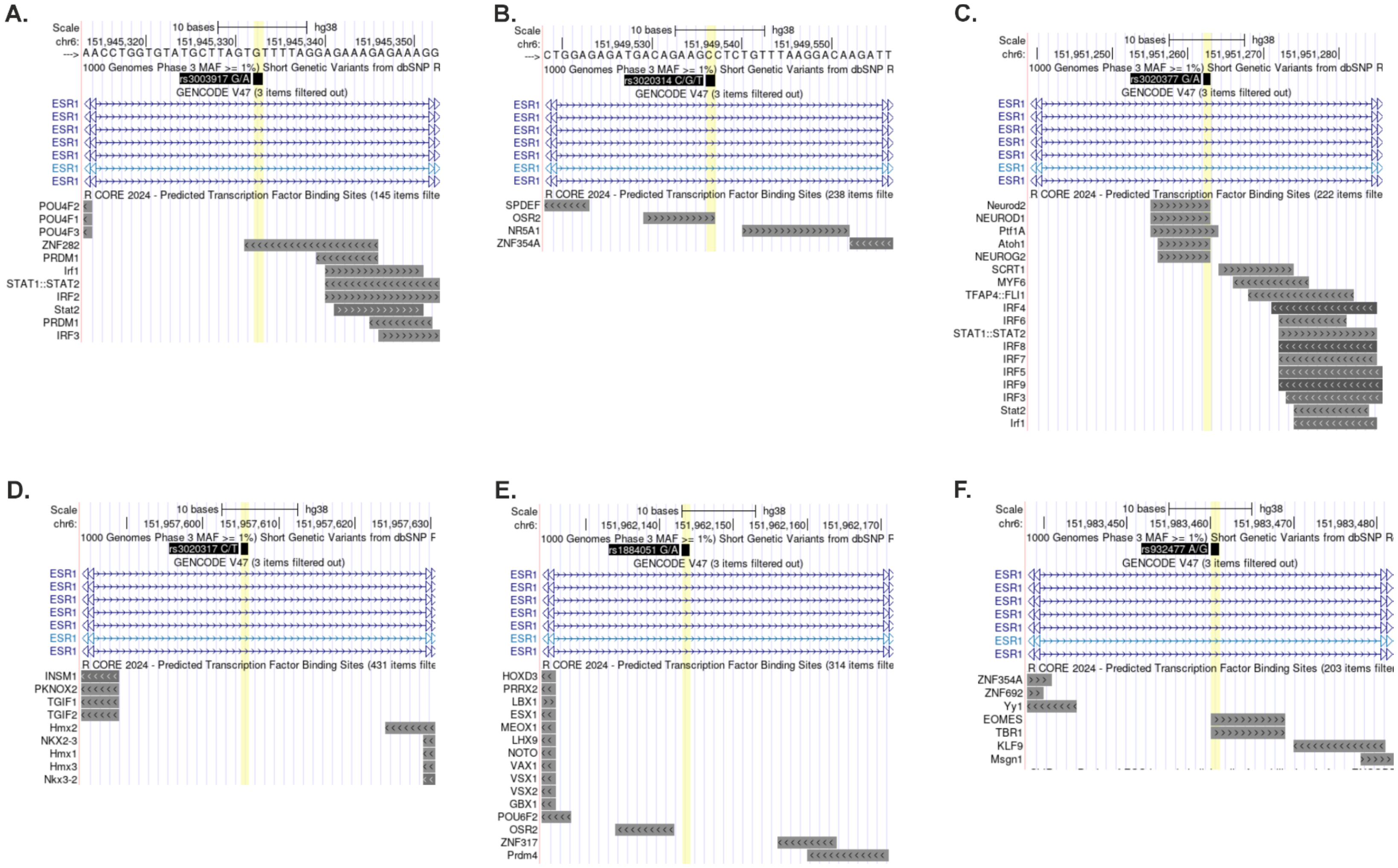

**Supplement Figure 2. Predicted transcription factor binding sites in the *ESR1* intron 4 region.** UCSC genome browser tracks show the six target SNPs (marked in yellow) **A.** rs3003917, **B.** rs3020314, **C.** rs3020377, **D.** rs3020317, **E.** rs1884051 and **F.** rs932477. All SNPs are directly overlapping with or at least in proximity (within max. 10-20 bp) to a predicted TF binding site.

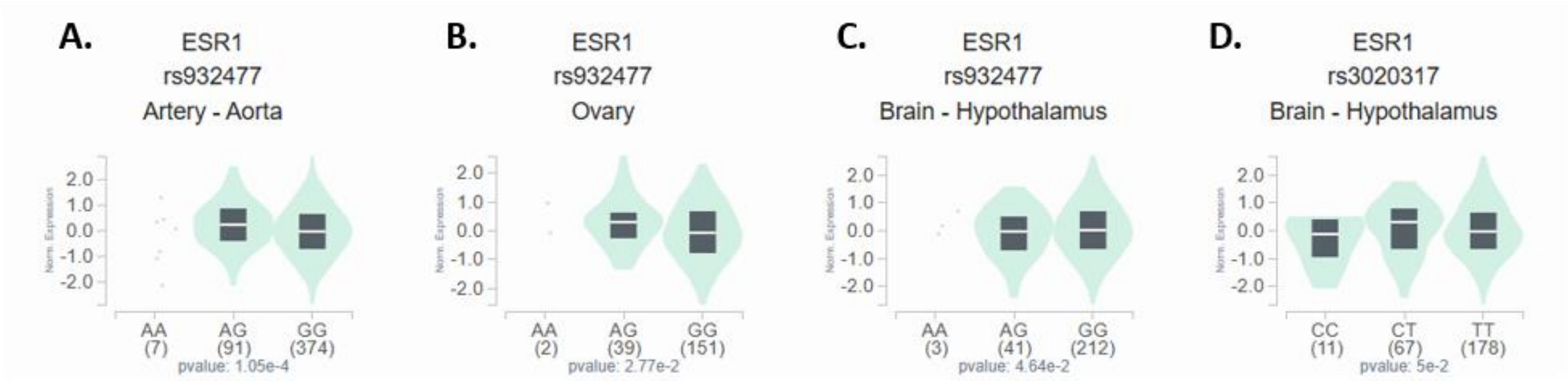

**Supplement Figure 3. Allele-dependent *ESR1* expression in different tissues based on GTEx data.** Significant influence on ESR1 expression was detected for rs932477 in **A.** Aorta, **B.** Ovary and **C.** Hypothalamus and for rs3020317 in **D.** Hypothalamus.

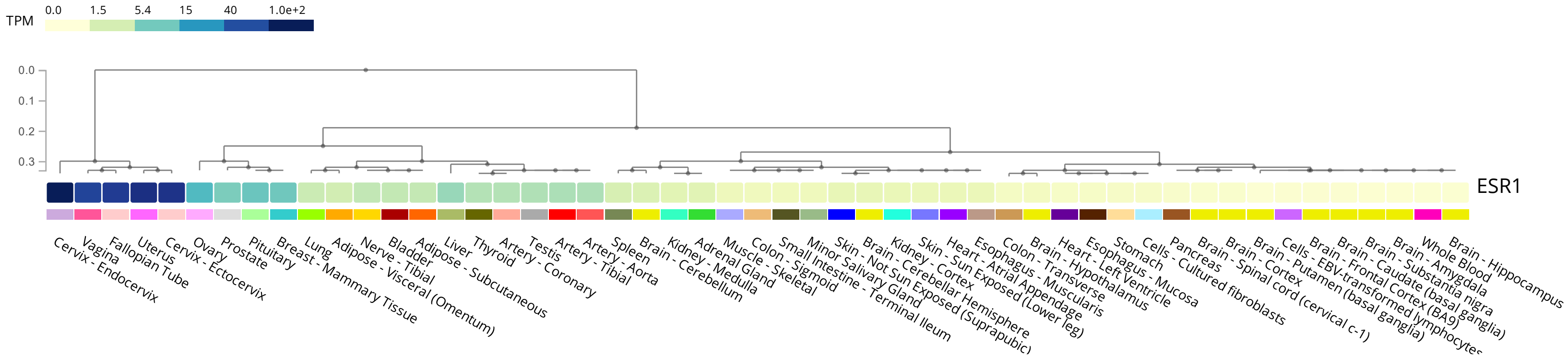

**Supplement Figure 4. ESR1 expression across different tissues based on GTEx data.** While ESR1 is either not expressed or expressed at very low levels in the brain (light yellow colors), its expression is higher in the breast (turquoise) and highest in the cervix (dark blue).

#### 3. Supplementary Tables

**Supplement Table 1. Analysis of Expression Quantitative Trait Loci (eQTL) of target SNPs in *ESR1*.**

| Gene | SNP | P-Value | Tissue |
|---|---|---|---|
| ESR1 | rs932477 | 0.00011 | Artery - Aorta |
| ESR1 | rs932477 | 0.028 | Ovary |
| ESR1 | rs932477 | 0.046 | Brain - Hypothalamus |
| ESR1 | rs3020317 | 0.050 | Brain - Hypothalamus |
| ESR1 | rs3020317 | 0.052 | Brain - Amygdala |
| ESR1 | rs932477 | 0.056 | Brain - Hippocampus |
| ESR1 | rs3003917 | 0.060 | Adipose - Subcutaneous |
| ESR1 | rs3003917 | 0.061 | Brain - Hypothalamus |
| ESR1 | rs3020377 | 0.066 | Adipose - Subcutaneous |
| ESR1 | rs3003917 | 0.067 | Brain - Cerebellum |
| ESR1 | rs3020314 | 0.068 | Adipose - Subcutaneous |
| ESR1 | rs3003917 | 0.069 | Ovary |
| ESR1 | rs932477 | 0.072 | Brain - Amygdala |
| ESR1 | rs3020317 | 0.078 | Brain - Cerebellum |
| ESR1 | rs3020377 | 0.080 | Brain - Hypothalamus |
| ESR1 | rs3020314 | 0.088 | Brain - Hypothalamus |
| ESR1 | rs3020377 | 0.10 | Brain - Cerebellum |
| ESR1 | rs1884051 | 0.10 | Brain - Hypothalamus |
| ESR1 | rs3020377 | 0.11 | Brain - Hippocampus |
| ESR1 | rs3020314 | 0.12 | Brain - Cerebellum |
| ESR1 | rs3020314 | 0.12 | Brain - Hippocampus |
| ESR1 | rs3003917 | 0.14 | Brain - Amygdala |
| ESR1 | rs1884051 | 0.17 | Brain - Cerebellum |
| ESR1 | rs3003917 | 0.18 | Brain - Hippocampus |
| ESR1 | rs3020317 | 0.19 | Adipose - Subcutaneous |
| ESR1 | rs3020314 | 0.21 | Lung |
| ESR1 | rs932477 | 0.21 | Muscle - Skeletal |
| ESR1 | rs3020377 | 0.22 | Lung |
| ESR1 | rs3003917 | 0.23 | Breast - Mammary Tissue |
| ESR1 | rs3020317 | 0.24 | Brain - Cortex |
| ESR1 | rs1884051 | 0.24 | Lung |
| ESR1 | rs3020317 | 0.25 | Breast - Mammary Tissue |
| ESR1 | rs1884051 | 0.28 | Brain - Hippocampus |
| ESR1 | rs1884051 | 0.39 | Adipose - Subcutaneous |
| ESR1 | rs3020377 | 0.39 | Artery - Aorta |
| ESR1 | rs3020317 | 0.39 | Heart - Atrial Appendage |
| ESR1 | rs3003917 | 0.39 | Liver |
| ESR1 | rs3020314 | 0.39 | Muscle - Skeletal |
| ESR1 | rs3020314 | 0.40 | Brain - Cortex |
| ESR1 | rs3003917 | 0.40 | Lung |
| ESR1 | rs3020377 | 0.40 | Muscle - Skeletal |
| ESR1 | rs3020317 | 0.41 | Ovary |

| | | | |
|---|---|---|---|
| ESR1 | rs3003917 | 0.42 | Brain - Cortex |
| ESR1 | rs932477 | 0.42 | Breast - Mammary Tissue |
| ESR1 | rs3020377 | 0.43 | Brain - Cortex |
| ESR1 | rs1884051 | 0.43 | Ovary |
| ESR1 | rs1884051 | 0.44 | Brain - Cortex |
| ESR1 | rs3020317 | 0.44 | Lung |
| ESR1 | rs3020314 | 0.45 | Artery - Aorta |
| ESR1 | rs3020317 | 0.46 | Artery - Aorta |
| ESR1 | rs932477 | 0.46 | Liver |
| ESR1 | rs3020314 | 0.48 | Ovary |
| ESR1 | rs932477 | 0.51 | Brain - Cerebellum |
| ESR1 | rs3020377 | 0.51 | Ovary |
| ESR1 | rs3020314 | 0.53 | Breast - Mammary Tissue |
| ESR1 | rs932477 | 0.54 | Brain - Cortex |
| ESR1 | rs3020317 | 0.56 | Liver |
| ESR1 | rs3020317 | 0.57 | Brain - Hippocampus |
| ESR1 | rs3020377 | 0.58 | Breast - Mammary Tissue |
| ESR1 | rs1884051 | 0.58 | Muscle - Skeletal |
| ESR1 | rs1884051 | 0.66 | Heart - Atrial Appendage |
| ESR1 | rs3020377 | 0.67 | Liver |
| ESR1 | rs1884051 | 0.69 | Artery - Aorta |
| ESR1 | rs3020314 | 0.69 | Liver |
| ESR1 | rs3020377 | 0.71 | Adrenal Gland |
| ESR1 | rs3003917 | 0.73 | Heart - Atrial Appendage |
| ESR1 | rs3020314 | 0.74 | Adrenal Gland |
| ESR1 | rs1884051 | 0.74 | Breast - Mammary Tissue |
| ESR1 | rs932477 | 0.79 | Adrenal Gland |
| ESR1 | rs3003917 | 0.80 | Artery - Aorta |
| ESR1 | rs3020377 | 0.80 | Brain - Amygdala |
| ESR1 | rs3020317 | 0.83 | Adrenal Gland |
| ESR1 | rs932477 | 0.83 | Lung |
| ESR1 | rs1884051 | 0.84 | Adrenal Gland |
| ESR1 | rs3020317 | 0.87 | Muscle - Skeletal |
| ESR1 | rs3020377 | 0.88 | Heart - Atrial Appendage |
| ESR1 | rs1884051 | 0.88 | Liver |
| ESR1 | rs1884051 | 0.89 | Brain - Amygdala |
| ESR1 | rs3020314 | 0.91 | Heart - Atrial Appendage |
| ESR1 | rs932477 | 0.93 | Heart - Atrial Appendage |
| ESR1 | rs3003917 | 0.95 | Adrenal Gland |
| ESR1 | rs932477 | 0.97 | Adipose - Subcutaneous |
| ESR1 | rs3020314 | 0.98 | Brain - Amygdala |
| ESR1 | rs3003917 | 0.99 | Muscle - Skeletal |
